# Emergency Manual Peritoneal Dialysis Using Adapted Nasogastric Tubes Versus Tenckhoff Catheters for Severe Acute Kidney Injury in Neonates and Infants: A Retrospective Cohort Study

**DOI:** 10.64898/2026.11.19.26368142

**Authors:** Ahror A. Alimov, Alisher M. Sharipov, Bahrom B. Mamatkulov, Suraye R. Rahimova

**Author notes:** **Corresponding Author:** Ahror A. Alimov, PhD, Department of Emergency Pediatrics, Tashkent State Medical University, 100109, Tashkent, Uzbekistan,.

## Abstract

**Background:** In neonates and young infants, extracorporeal renal replacement therapy (RRT) for acute kidney injury (AKI) is often not feasible, and peritoneal dialysis (PD) remains the most practical option. In resource-limited settings, however, dedicated pediatric PD catheters are frequently unavailable.

**Objective:** To evaluate the efficacy and safety of emergency manual PD in critically ill neonates and infants with severe AKI, and to compare complications between adapted nasogastric tubes and Tenckhoff catheters.

**Methods:** In this retrospective cohort study, 28 critically ill children aged up to 3 months (16 boys and 12 girls) received emergency PD between January 2022 and June 2023. Access was obtained with a Tenckhoff catheter in 8 patients (28.6%) and an adapted 12 Fr nasogastric tube in 20 (71.4%). Renal, metabolic, fluid balance and hemodynamic (Vasoactive-Inotropic Score, VIS) parameters were assessed before and during PD.

**Results:** AKI followed cardiac surgery with cardiopulmonary bypass in 10 patients (35.7%), sepsis in 12 (42.9%), and congenital pneumonia after complicated delivery in 6 (21.4%). Serum urea, creatinine and potassium decreased significantly (all p < 0.001). Within 72 hours, the daily fluid balance shifted from +42.3 ± 14.8 to −12.4 ± 8.6 mL/kg/day, and the mean VIS fell from 40.4 ± 9.6 to 5.3 ± 3.1 (both p < 0.001). Complication rates did not differ significantly between access types, and no peritonitis occurred. All patients recovered renal function, and there were no deaths.

**Conclusions:** In this small retrospective cohort, emergency manual PD was associated with rapid metabolic correction, reversal of fluid overload and hemodynamic improvement. An adapted nasogastric tube may be a feasible temporary alternative for peritoneal access when Tenckhoff catheters are unavailable; prospective studies are needed to confirm its safety.

## Introduction

Acute kidney injury (AKI) is a common and serious complication in critically ill neonates and young infants, particularly in those undergoing cardiac surgery for congenital heart disease and those with sepsis or severe perinatal respiratory disease. In the multinational AWAKEN cohort, AKI occurred in approximately 30% of neonates admitted to intensive care and was independently associated with higher mortality and longer hospital stay ^1^. Where renal replacement therapy (RRT) is required but unavailable, as is often the case in peripheral and resource-limited regions, outcomes are considerably worse ^2^. In a survey of 35 European pediatric nephrology centers, peritoneal dialysis (PD) and continuous RRT (CRRT) were the most frequently used modalities (39.4% and 38.2% of treatments, respectively), followed by intermittent hemodialysis (HD; 22.4%) ^3^, and PD was the preferred modality in centers with cardiac surgery programs ^4^.

In newborns, extracorporeal therapies are limited by large circuit volumes relative to blood volume, the need for anticoagulation, and difficult vascular access. Consequently, PD remains the most widely used form of RRT in neonates and young infants, owing to its relative ease of access and technical simplicity ^5,6^. Acute PD can be initiated in units without hemodialysis expertise and is effective in correcting AKI-related fluid and metabolic disturbances in children of all ages, including neonates and preterm infants ^7^.

Automated PD cyclers have improved the safety of PD; however, they cannot reliably deliver the very small exchange volumes required by neonates and small infants. In this population, PD is usually performed manually using closed exchange systems, which are inexpensive and well suited to very young patients ^8–10^. Moreover, dedicated pediatric PD catheters are frequently unavailable in emergency and low-resource settings, and clinicians have successfully improvised peritoneal access with alternative devices, including urinary (Foley) catheters, nasogastric tubes and suprapubic catheters, with acceptable outcomes and reduced costs ^11^.

However, data on the safety of such improvised access in critically ill neonates and infants, and on the hemodynamic effects of early PD in this population, remain scarce. The aim of this study was therefore to evaluate the efficacy and safety of emergency manual PD in critically ill neonates and infants with severe AKI, and to compare access-related complications between adapted nasogastric tubes and standard Tenckhoff catheters.

## Methods

### Study Design and Population

We conducted a retrospective cohort study of critically ill neonates and infants who underwent emergency PD for severe AKI between January 2022 and June 2023 at the National Children’s Medical Center (NCMC), Tashkent, and in the neonatal intensive care units (NICUs) of regional children’s hospitals. The cohort comprised 28 patients aged up to 3 months: 12 neonates (≤ 28 days of age) and 16 infants. All patients were born at term. AKI was diagnosed and staged according to the neonatal modified Kidney Disease: Improving Global Outcomes (KDIGO) criteria ^1^. Renal Doppler ultrasonography was performed in all patients before RRT to exclude post-renal (obstructive) causes. The study is reported in accordance with the STROBE guidelines for observational studies. Patients were enrolled consecutively, and no exclusion criteria were applied.

### Indications and PD Initiation

The decision to initiate acute PD was made when fluid overload and/or electrolyte and acid– base disturbances remained refractory to at least 12 hours of conservative management. Indications were: (1) severe oliguria or anuria (urine output < 0.5 mL/kg/h) unresponsive to fluid resuscitation and high-dose loop diuretics; (2) clinically significant fluid overload, with a markedly positive daily fluid balance and generalized edema; and (3) persistent severe metabolic acidosis.

### Catheter Placement and PD Procedure

In 8 patients treated at the NCMC, a straight Tenckhoff catheter was inserted surgically by a pediatric surgeon under general anesthesia. In the remaining 20 patients, mostly treated in regional hospitals where pediatric PD catheters were unavailable and hemodynamic instability precluded safe transfer, a sterile 12 Fr nasogastric tube was adapted for peritoneal access. The tube was inserted through a semi-open approach: a small incision was made in the left lower abdominal quadrant and dissection was carried down to the rectus sheath, after which the tube was advanced into the pelvic cavity using a modified Seldinger technique. The exit site was sutured and covered with a sterile occlusive dressing (Figure 1).

**Figure 1.**
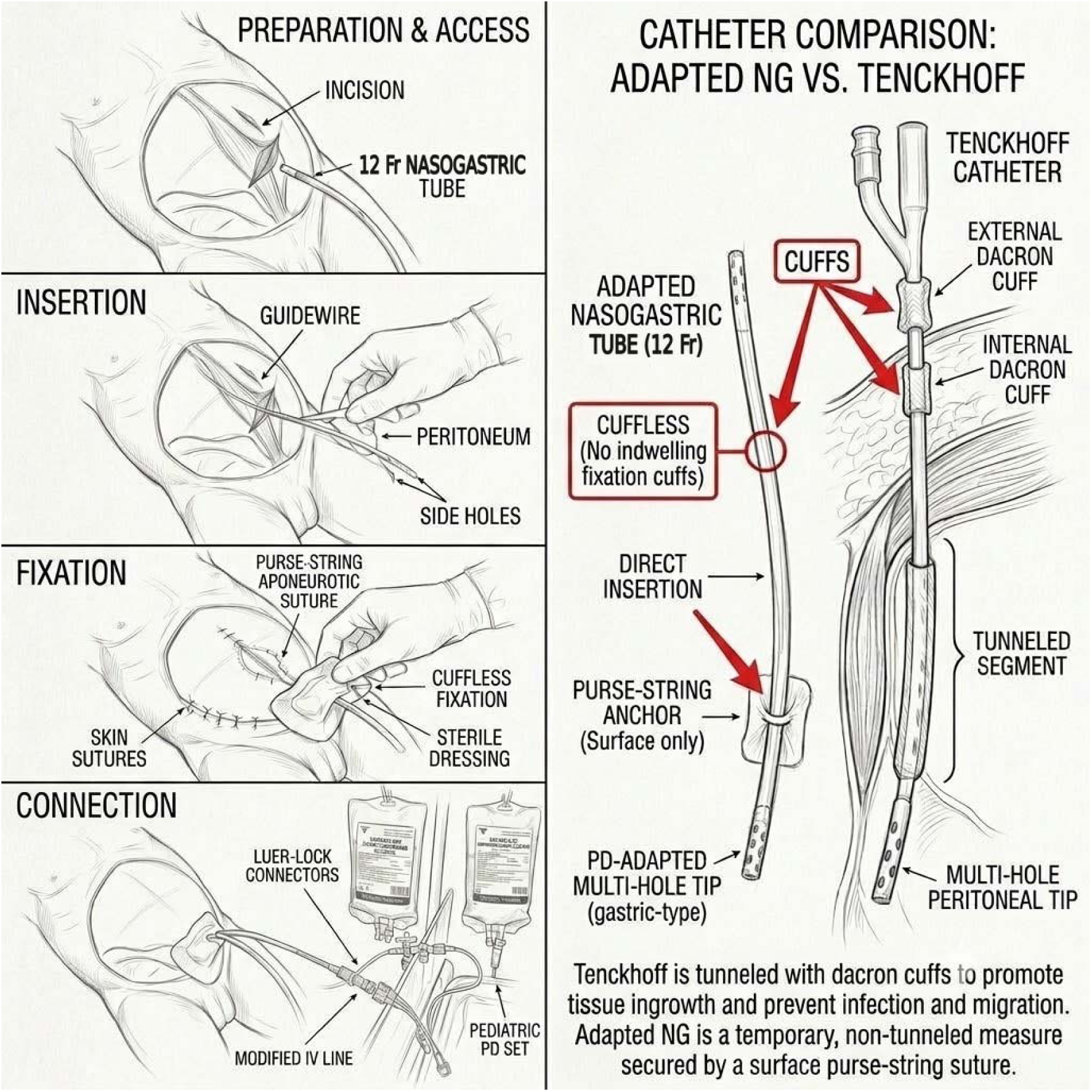
Schematic representation of the adapted 12 Fr nasogastric tube inserted for peritoneal dialysis and its structural comparison with a standard Tenckhoff catheter.

PD was performed manually using a closed system assembled from standard intravenous infusion lines. Exchange volumes started at 5–10 mL/kg and were gradually increased to 15 mL/kg as tolerated. Bicarbonate-buffered dialysate with dextrose concentrations of approximately 1.5%, 2.5%, or 4.5% was prepared from standard intravenous solutions (Table 1) and selected according to the required ultrafiltration and serum glucose levels. Dwell times were initially approximately 70 minutes and were extended to 2–3 hours after exit-site healing. Heparin (500 U/L) was added to the dialysate during the first 48 hours, until the effluent was free of blood. A first-generation cephalosporin (cefazolin, 100 mg/kg/day) was administered intravenously as prophylaxis before catheter insertion. Serum glucose, calcium and potassium were monitored by arterial blood gas analysis (ABL 8000).

**Table 1.**
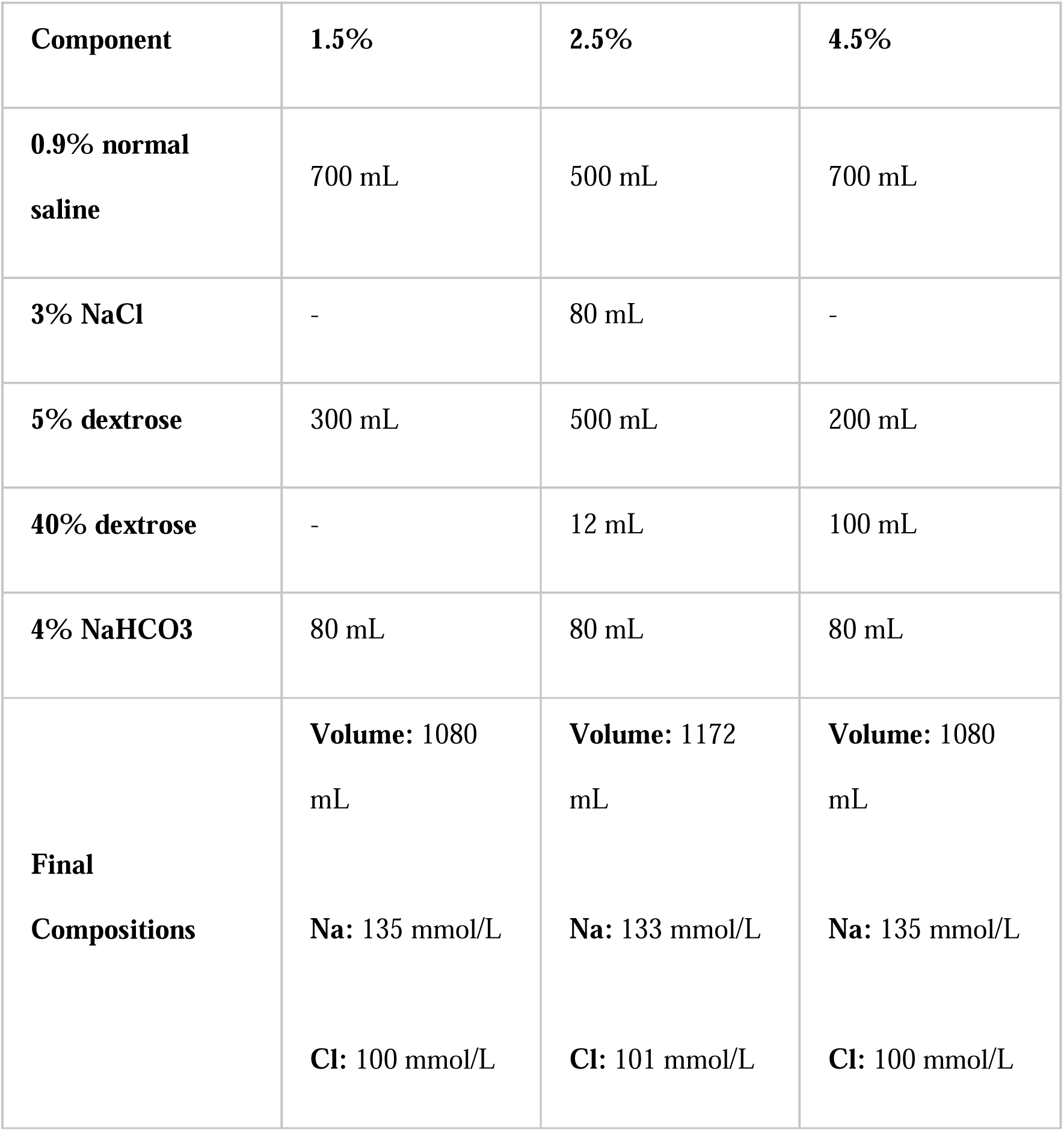
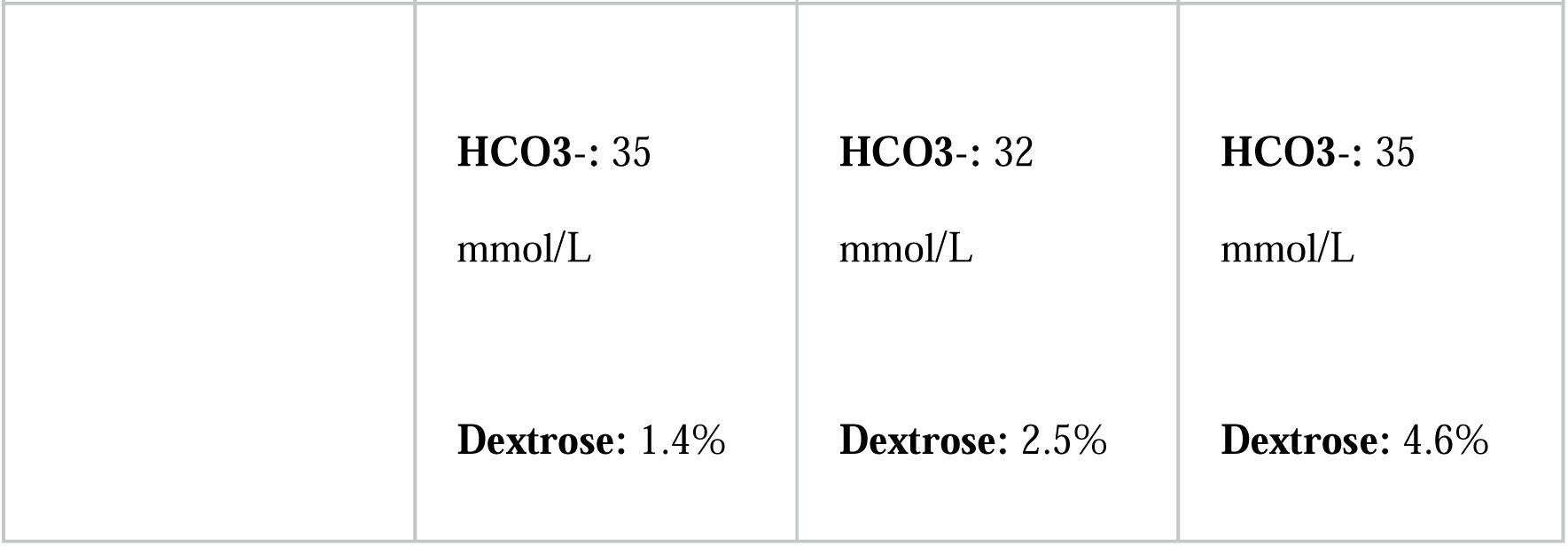
Composition of the locally prepared dialysate solutions^12^.

| **Component** | **1.5%** | **2.5%** | **4.5%** |
|---|---|---|---|
| **0.9% normal saline** | 700 mL | 500 mL | 700 mL |
| **3% NaCl** | - | 80 mL | - |
| **5% dextrose** | 300 mL | 500 mL | 200 mL |
| **40% dextrose** | - | 12 mL | 100 mL |
| **4% NaHCO3** | 80 mL | 80 mL | 80 mL |
| **Final Compositions** | **Volume:** 1080 mL<br>**Na:** 135 mmol/L<br>**Cl:** 100 mmol/L<br>**HCO3**-**:** 35 mmol/L<br>**Dextrose:** 1.4% | **Volume:** 1172 mL<br>**Na:** 133 mmol/L<br>**Cl:** 101 mmol/L<br>**HCO3**-**:** 32 mmol/L<br>**Dextrose:** 2.5% | **Volume:** 1080 mL<br>**Na:** 135 mmol/L<br>**Cl:** 100 mmol/L<br>**HCO3**-**:** 35 mmol/L<br>**Dextrose:** 4.6% |

### Data Collection and Clinical Definitions

Medical records were reviewed to extract demographic data, the primary etiology of AKI, type of peritoneal access, laboratory values, clinical outcomes, and PD-related complications. Hemodynamic support was quantified using the Vasoactive-Inotropic Score (VIS) at baseline and after 24 and 72 hours of PD, calculated as: VIS = dopamine + dobutamine + 100 × epinephrine + 100 × norepinephrine + 10 × milrinone (all in µg/kg/min) + 10,000 × vasopressin (U/kg/min) ^13^. Fluid intake, output (urine and PD ultrafiltration) and daily fluid balance were recorded. Renal recovery was defined as restoration of urine output together with a progressive decline in serum urea and creatinine and regression of pathological changes on renal ultrasonography, allowing PD to be discontinued. Patients were followed throughout the course of PD until its discontinuation (maximum 14 days). Baseline refers to the time of PD initiation. Daily fluid balance was calculated as total fluid intake minus total output (urine and net PD ultrafiltration). Mean vasoactive doses and VIS were calculated for all 28 patients, with patients not receiving vasoactive drugs scored as zero.

### Statistical Analysis

Given the small sample size, the analysis was primarily descriptive. Continuous variables are presented as mean ± standard deviation (SD) or median [interquartile range, IQR], according to their distribution. Categorical variables are expressed as n (%). Complication rates between access types were compared using Fisher’s exact test. Pre- and post-PD laboratory values were compared using the paired Student’s t-test or the Wilcoxon signed-rank test, as appropriate. Variables measured at baseline, 24 h and 72 h (VIS, mean arterial pressure, fluid intake and fluid balance) were analyzed with the Friedman test, and the proportion of patients requiring vasoactive support with Cochran’s Q test. A two-sided p < 0.05 was considered statistically significant. Data were collected in Microsoft Excel and Access; statistical analyses were performed using IBM SPSS Statistics (IBM Corp., Armonk, NY, USA).

## Results

### Baseline Characteristics

A total of 28 patients were included: 12 neonates and 16 infants (Table 2). The median age of the neonates was 7 days (IQR 4–12) and their mean body weight 3.4 ± 0.6 kg; the median age of the infants was 62 days (IQR 38–120) and their mean weight 5.8 ± 1.4 kg. The etiology of AKI differed between the two age groups: in neonates, the predominant cause was cardiac surgery with prolonged cardiopulmonary bypass (CPB) (83.3%), whereas in infants AKI was mainly associated with sepsis (62.5%) and congenital pneumonia after complicated delivery (37.5%). At baseline, 50% of patients required mechanical ventilation and 57.1% required vasoactive support. A Tenckhoff catheter was used in 8 patients (28.6%) and an adapted 12 Fr nasogastric tube in 20 patients (71.4%). The duration of PD ranged from 3 to 14 days.

**Table 2.** Baseline demographic and clinical characteristics of the patients.

| **Parameter** | **Neonates (n=12)** | **Infants (n=16)** | **Total (n=28)** |
|---|---|---|---|
| **Age (days)**, median [IQR] | 7 [4; 12] | 62 [38; 120] | 18 [6; 68] |
| **Body weight (kg)**, mean ± SD | 3.4 ± 0.6 | 5.8 ± 1.4 | 4.8 ± 1.6 |
| **Sex (Male / Female)** | 7 / 5 | 9 / 7 | 16 / 12 |
| **Etiology of AKI, n (%)** |  |  |  |
| Post-cardiac surgery (CPB) | 10 (83.3%) | 0 (0%) | 10 (35.7%) |
| Sepsis / Septicemia | 2 (16.7%) | 10 (62.5%) | 12 (42.9%) |
| Complicated delivery / congenital pneumonia | 0 (0%) | 6 (37.5%) | 6 (21.4%) |
| **Peritoneal access type, n (%)** |  |  |  |
| Tenckhoff catheter | 3 (25.0%) | 5 (31.3%) | 8 (28.6%) |
| Nasogastric tube (12 Fr) | 9 (75.0%) | 11 (68.8%) | 20 (71.4%) |
| **Mechanical ventilation**, n (%) | 9 (75.0%) | 5 (31.3%) | 14 (50.0%) |
| **Vasopressor requirement**, n (%) | 8 (66.7%) | 8 (50.0%) | 16 (57.1%) |

### Renal and Metabolic Outcomes

PD was associated with rapid and significant improvement in renal and metabolic parameters (Table 3). Mean serum urea decreased from 14.2 ± 4.1 mmol/L at baseline to 7.8 ± 2.6 mmol/L at the end of PD (p < 0.001), and serum creatinine from 187.3 ± 52.4 µmol/L to 98.6 ± 31.7 µmol/L (p < 0.001). Serum sodium remained stable (138.4 ± 6.2 vs. 137.1 ± 4.8 mmol/L, p = 0.24), whereas serum potassium decreased from 5.8 ± 1.1 to 4.4 ± 0.7 mmol/L (p < 0.001). Urine output increased from < 0.5 mL/kg/h at baseline to 1.2 ± 0.5 mL/kg/h (p < 0.001).

**Table 3.** Dynamics of clinical and laboratory parameters during peritoneal dialysis (n=28)

| **Parameter** | **Baseline (Pre-PD)** | **End of PD** | **p-value** |
|---|---|---|---|
| **Serum Urea (mmol/L)** | 14.2 ± 4.1 | 7.8 ± 2.6 | < 0.001 |
| **Serum Creatinine (µmol/L)** | 187.3 ± 52.4 | 98.6 ± 31.7 | < 0.001 |
| **Sodium (mmol/L)** | 138.4 ± 6.2 | 137.1 ± 4.8 | 0.24 |
| **Potassium (mmol/L)** | 5.8 ± 1.1 | 4.4 ± 0.7 | < 0.001 |
| **Urine output (mL/kg/h)** | < 0.5 | 1.2 ± 0.5 | < 0.001 |

### Fluid Balance and Hemodynamics

Early PD was accompanied by resolution of fluid overload and stabilization of systemic hemodynamics (Table 4). Before PD, patients had a mean positive fluid balance of +42.3 ± 14.8 mL/kg/day and a mean VIS of 40.4 ± 9.6. After 72 hours of PD, with ultrafiltration averaging 31.4 ± 10.6 mL/kg/day, the daily fluid balance had become negative (−12.4 ± 8.6 mL/kg/day; p < 0.001).

**Table 4.** Hemodynamic and fluid balance dynamics during early peritoneal dialysis (n=28)

| **Parameter** | **Baseline (Pre-PD)** | **24h of PD** | **72h of PD** | **p-value (overall)** |
|---|---|---|---|---|
| **Mean Arterial Pressure (mmHg)** | 48.2 ± 9.4 | 52.6 ± 8.1 | 61.3 ± 7.2 | < 0.001 |
| **Vasoactive-Inotropic Score (VIS)** | 40.4 ± 9.6 | 22.4 ± 7.1 | 5.3 ± 3.1 | < 0.001 |
| **Patients requiring vasopressors, n (%)** | 16 (57.1%) | 14 (50.0%) | 6 (21.4%) | 0.004 |
| **Total fluid intake (mL/kg/day)** | 148.4 ± 22.6 | 138.2 ± 19.4 | 121.6 ± 16.8 | < 0.001 |
| **PD Ultrafiltration volume (mL/kg/day)** | — | 24.8 ± 9.2 | 31.4 ± 10.6 | — |
| **Daily fluid balance (mL/kg/day)** | +42.3 ± 14.8 | +18.6 ± 11.2 | −12.4 ± 8.6 | < 0.001 |

In parallel, mean arterial pressure (MAP) increased from 48.2 ± 9.4 to 61.3 ± 7.2 mmHg (p < 0.001), and the mean VIS decreased to 5.3 ± 3.1 (p < 0.001). Norepinephrine was discontinued in all patients, and the proportion of patients requiring vasoactive support decreased from 57.1% to 21.4% (p = 0.004).

### Complications and Outcomes by Access Type

Access-related complication rates did not differ significantly between the two catheter types, although the study was not powered to detect such differences (Table 5). Dialysate leakage occurred in 2 of 8 patients (25.0%) with Tenckhoff catheters and in 3 of 20 (15.0%) with nasogastric tubes (p = 0.61). Catheter obstruction or kinking (12.5% vs. 20.0%, p > 0.99) and exit-site infection (12.5% vs. 5.0%, p = 0.50) were also comparable and were managed conservatively or by catheter replacement. No episodes of peritonitis occurred in either group.

**Table 5.** Complications and clinical outcomes according to catheter type.

| **Outcomes and Complications** | **Tenckhoff (n=8)** | **Nasogastric Tube (n=20)** | **p-value** |
|---|---|---|---|
| **Dialysate leakage, n (%)** | 2 (25.0%) | 3 (15.0%) | 0.61 |
| **Catheter obstruction / kinking, n (%)** | 1 (12.5%) | 4 (20.0%) | > 0.99 |
| **Exit-site infection, n (%)** | 1 (12.5%) | 1 (5.0%) | 0.50 |
| **Peritonitis, n (%)** | 0 (0%) | 0 (0%) | > 0.99 |
| **Recovery of renal function, n (%)** | 8 (100%) | 20 (100%) | > 0.99 |
| **Mortality, n (%)** | 0 (0%) | 0 (0%) | > 0.99 |

Renal function recovered in all 28 patients, and there were no deaths during PD and until hospital discharge.

## Discussion

In critically ill neonates and young children, AKI is an independent predictor of mortality, with reported mortality rates of up to 45% among those requiring RRT in intensive care ^14–17^. In our cohort of 28 critically ill neonates and infants treated with emergency PD, all patients recovered renal function and none died. Although these outcomes compare favorably with published series^14–17^, they should be interpreted with caution given the small sample, the absence of a control group, and possible selection bias. Nevertheless, our findings support the feasibility and effectiveness of timely PD in oligoanuric AKI when conservative management fails.

A key observation of this study was the improvement in systemic hemodynamics that accompanied correction of fluid overload. Most patients presented with marked cardiovascular compromise, due predominantly to low cardiac output after cardiac surgery in neonates and to septic shock in infants, and required substantial vasoactive support. Over 72 hours, PD ultrafiltration shifted the daily fluid balance from markedly positive (+42.3 ± 14.8 mL/kg/day) to negative (−12.4 ± 8.6 mL/kg/day), accompanied by an increase in mean arterial pressure and a significant reduction in VIS from 40.4 to 5.3. These data suggest that early PD may act not only as a renal replacement modality but also as an adjunct to hemodynamic stabilization, potentially interrupting the cycle of fluid overload, myocardial and tissue edema, and escalating vasoactive requirements. However, because concomitant treatment (antimicrobial therapy and source control in sepsis, and postoperative recovery after cardiac surgery) also contributed to recovery, a causal relationship cannot be established from our data.

Reluctance to use PD in critically ill infants often stems from concerns about procedure-related complications, such as peritonitis, dialysate leakage and catheter obstruction ^18,19^. In our cohort, the adapted nasogastric tube was used in 71.4% of cases because of resource constraints and the inability to transfer hemodynamically unstable patients. Complication rates did not differ significantly between nasogastric tubes and Tenckhoff catheters, and no cases of peritonitis occurred. Given the small groups, however, the absence of a statistically significant difference cannot be interpreted as equivalence. The absence of peritonitis may reflect periprocedural antibiotic prophylaxis, strict aseptic technique and the short duration of dialysis, although our study was not designed to assess the contribution of each factor.

Overall, our findings suggest that an adapted nasogastric tube is a feasible, low-cost temporary option for acute PD access when dedicated pediatric catheters are unavailable, consistent with previous reports of improvised access devices ^11,18^. Our initial exchange volumes (5–10 mL/kg) were also lower than those suggested by the ISPD 2020 paediatric guideline (initial fill of 10–20 mL/kg, increasing to about 30–40 mL/kg)^2^, and an uncuffed, non-tunneled tube was used in most patients while PD lasted up to 14 days, whereas the guideline considers a surgically placed Tenckhoff catheter the optimal access^2^. These departures from guideline practice should be kept in mind when interpreting our results.

Serum urea and potassium decreased rapidly after PD initiation. Serum creatinine also fell significantly, although more gradually, consistent with previous reports ^20,21^; this is expected, because peritoneal clearance of creatinine is slower than that of urea and the fall in creatinine also reflects recovery of endogenous renal function. Because it requires neither vascular access nor anticoagulation and has minimal hemodynamic impact, manual PD remains a practical RRT option for neonates and small infants in peripheral or resource-limited settings ^22,23^.

### Limitations

This study has several limitations. First, it was retrospective and included a small number of patients without a control group, which precludes causal inference and limits statistical power, particularly for the comparison of complication rates between access types. Second, the access device was not randomly assigned but determined by the treating center and device availability; Tenckhoff catheters were placed mainly at the tertiary center and nasogastric tubes mainly in regional hospitals, so differences in staff experience and care may have confounded this comparison. Third, the use of a nasogastric tube for peritoneal access is an off-label adaptation performed as an emergency measure; our data address only short-term use, and because follow-up was limited to the dialysis period (maximum 14 days), long-term renal outcomes were not assessed. Fourth, only term infants were included, so the findings cannot be extrapolated to preterm or low-birth-weight neonates. Prospective multicenter studies are needed to establish standardized protocols for alternative peritoneal access in emergency neonatal PD. Fifth, the dialysate was prepared locally from intravenous solutions and contained no calcium, magnesium or potassium (Table 1) by biochemical analysis. Finally, the fall in VIS may partly reflect the natural postoperative course and the treatment of sepsis, and no control group was available to separate these effects.

## Conclusions

In this retrospective cohort of critically ill neonates and infants with severe AKI, emergency manual PD was associated with rapid correction of azotemia and hyperkalemia, reversal of fluid overload and a marked reduction in vasoactive support, with full renal recovery and no mortality. When dedicated pediatric catheters are unavailable, an adapted nasogastric tube may be a feasible temporary alternative for peritoneal access. These findings warrant confirmation in larger prospective studies.

## Supporting information

Titlepage

Title Article

## Ethics Approval and Consent to Participate

The study was approved by the Local Ethics Committee of the National Children’s Medical Center, Tashkent, Uzbekistan (Protocol No. 1, 20 January 2025). The committee approved the use of retrospectively collected, anonymized clinical data for scientific publication. No additional diagnostic or therapeutic interventions were performed for research purposes, and the study was conducted in accordance with the principles of the Declaration of Helsinki. Committee decision covers the regional hospitals.

## Consent for Publication

This manuscript contains no photographs or other details that could identify an individual patient; only aggregated, anonymized data are reported.

Informed consent for the use of anonymized patient data was obtained through the ethics committee.

## Declaration of Generative AI and AI-Assisted Technologies

During the preparation of this manuscript, the authors used Google Gemini and Claude (Anthropic) for translation, language editing and table formatting. After using these tools, the authors reviewed and edited the content as needed and take full responsibility for the content of the publication. Figure 1 was edited using Google Gemini to enhance and structure the schematic of the procedure.

## Author Contributions

A.A.A. (Ahror A. Alimov): conceptualization, methodology, investigation, data curation, formal analysis, writing – original draft. A.M.S. (Alisher M. Sharipov): supervision, project administration, writing – review and editing. B.B.M. (Bahrom B. Mamatkulov): clinical (nephrology) consultation, validation, writing – review and editing. S.R.R. (Suraye R. Rahimova): clinical (critical care) consultation, validation, writing – review and editing. All authors approved the final manuscript.

## Data Availability Statement

The data supporting the findings of this study are available from the corresponding author upon reasonable request.

