## Supplementary material for "Emergency Manual Peritoneal Dialysis Using Adapted Nasogastric Tubes Versus Tenckhoff Catheters for Severe Acute Kidney Injury in Neonates and Infants: A Retrospective Cohort Study": Titlepage

**Title page**

Running title: Peritoneal Dialysis for Neonatal AKI

**Authors**

Ahror A. Alimov, PhD ¹ (ORCID 0000-0001-6430-3295)

Alisher M. Sharipov, DSc ¹,² (ORCID 0009-0002-9014-6793)

Bahrom B. Mamatkulov, PhD ² (ORCID 0000-0003-1921-4458)

Suraye R. Rahimova, MD ³ (ORCID 0009-0005-9856-2590)

**Affiliations**

¹ Department of Emergency Pediatrics, Tashkent State Medical University, 100109, Tashkent, Uzbekistan

² National Children's Medical Center postal code:100054, Tashkent, Uzbekistan

³ Department of Emergency Pediatrics, Republican Research Centre of Emergency Medicine, 100115, Tashkent, Uzbekistan

**Corresponding author**

Ahror A. Alimov, PhD, Department of Emergency Pediatrics, Tashkent State Medical University, 100109, Tashkent, Uzbekistan.

**Declarations**

Ethics approval: The study was approved by the Local Ethics Committee of the National Children's Medical Center, Tashkent, Uzbekistan (Protocol No. 1, 20 January 2025). The study was conducted in accordance with the Declaration of Helsinki. Committee decision covers the regional hospitals. Consent for publication: This manuscript contains no photographs or other details that could identify an individual patient; only aggregated, anonymized data are reported. Informed consent for the use of anonymized patient data was obtained through the ethics committee.

Data availability: The data supporting the findings of this study are available from the corresponding author upon reasonable request.

Conflict of interest: The authors declare no conflicts of interest.

Funding: The authors received no specific funding for this study.

Use of generative AI: During the preparation of this work, the authors used Google Gemini and Claude (Anthropic) for translation, language editing and table formatting. Figure 1 was edited using Google Gemini to enhance and structure the schematic of the procedure. The authors reviewed and edited the content and take full responsibility for the manuscript.
